# A Systematic Review on Medical Oxygen Ecosystem: Current State and Recent Advancements

**DOI:** 10.1101/2022.10.23.22281394

**Authors:** Ehtashamul Haque, Saber Al Tarek, Farhana Sarker, Md. Atiqul Haque, Khondaker A. Mamun

**Affiliations:** Advanced Intelligent Multidisciplinary Systems Lab (AIMS Lab), Institute of Research, Innovation, Incubation and Commercialization (IRIIC), United International University (UIU), Dhaka 1212, Bangladesh; CMED Health Limited, Dhaka 1206, Bangladesh; Department of Public Health & Informatics, Bangabandhu Sheikh Mujib Medical University, Dhaka 1000, Bangladesh; Department of Computer Science and Engineering, United International University (UIU), Dhaka 1212, Bangladesh

## Abstract

**Background:** Medical oxygen is an essential component of modern healthcare, with a wide variety of applications ranging from supplemental use in surgery and trauma patients to the primary medication in oxygen therapy. This is the most effective treatment for any respiratory illness. Despite the importance of oxygen for public health and its demand as a life-saving drug, research on the subject is limited, with the majority of studies conducted following the outbreak of the COVID-19 pandemic. Due to the lack of empirical studies, we aimed to compile the recent research efforts with the current state of the field through a systematic review.

**Methods:** We have performed a systematic review targeting the medical oxygen ecosystem, following the Preferred Reporting Items for Systematic review and Meta-Analysis Protocols (PRISMA-P). For the study, we have limited our scope to healthcare facilities and domiciliary applications of medical oxygen. We considered the articles published in the last twenty years, starting from the SARS outbreak in November 2002 to 15^th^ May 2022.

**Results:** Our systematic search resulted in forty-one preliminary articles, with three more articles appended for a complete outlook on the topic. Based on the selected articles, the current state of the topic was presented through detailed discussion and analysis.

**Conclusion:** We have presented an in-depth discussion of the research works found through the systematic search while extrapolating to provide insights on the current subject scenario. We have highlighted the areas with inadequate contemporary studies and presented some research gaps in the field.

## Introduction

Oxygen is an essential supplement in most medical procedures, as well as a prescribed treatment for different diseases related to the respiratory tract. Oxygen therapy is the most common treatment for respiratory distress, where medical-grade oxygen is provided to the patient to alleviate the symptoms [1]. Oxygen therapy is required for patients with conditions including, but not limited to, acute trauma to the respiratory system, decompression sickness, chronic obstructive pulmonary disease (COPD), pneumonia, asthma, and recently COVID-19.

According to WHO guidelines, medicine-grade oxygen must contain at least 82% pure oxygen and be produced using an oil-free compressor without contamination [2]. All over the world, the composition of medical oxygen is closely regulated by the national agencies of every country. Although the main form of oxygen generation is the same for medical and industrial oxygen, industrial-grade oxygen is allowed to have more impurities compared to medical oxygen and isn’t as firmly regulated [3]. We are restricting the scope of our study only to clinical oxygen.

Research relating to medical oxygen has mostly been neglected until the recent emergence of the COVID-19 pandemic, when the world faced an acute shortage of oxygen [4]. We believe the lack of systematic exploration in this field prior to the COVID-19 pandemic contributed to the exacerbation of the crisis encountered during the outbreak. Our goal for this study is to present a systematic review of the medical oxygen ecosystem following the Preferred Reporting Items for Systematic review and Meta-Analysis Protocols (PRISMA-P) [5]. This work highlights the current state and advances in the last 20 years of oxygen systems in hospital and home settings and identifies the research gaps and current trends in this domain.

## Methods

The systematic review was conducted adhering to the PRISMA-P checklist. The checklist is provided as an appendix (see Appendix 1).

### Eligibility criteria

As our study tries to compile recent advancements, we have considered a period of 20 years starting from the outbreak of SARS (Severe acute respiratory distress) in November 2002 [6]. We limited our scope to only medicine-grade oxygen as per WHO guidelines [2]. Only articles in English language were selected and we limited the type of documents to journal or conference articles and editorial letters for the preliminary search. Additional articles were added only when absolutely needed. Studies not performed in or for a hospital or home setting were rejected.

### Search strategy

Using the inclusion and exclusion criteria, we, at first, came up with the following search string - *(“medical oxygen” OR “hospital oxygen” OR “clinical oxygen”) AND system* which had returned very limited amount of search results. Hence, to increase the number of search results, all the possible fields in a medical oxygen ecosystem had been added, which resulted in the final search string to be - *(“medical oxygen” OR “hospital oxygen” OR “clinical oxygen”) AND (planning OR produc* OR generat* OR storage OR optim* OR demand OR supply OR monitor* OR manag* OR distribution OR protection OR system OR adminis*)*. For our systematic review, we have considered six different databases which are as follows -

1. Scopus
2. PubMed
3. SAGE
4. IEEE Xplore
5. Springer
6. Wiley

All the searches were made on 15^th^ May 2022. The exact search strings for each search engine are provided as an appendix (see Appendix 2).

### Selection procedure

We carried out the article screening following the standard PRISMA framework. While searching the articles, we exported the results with their abstracts in comma-separated values (CSV) format. Afterward, we appended all the records from different databases into a single file and removed the duplicates. We performed the 1^st^ screening on the unique articles by going through their titles and abstracts. During this screening, we dropped an article only if it met all the exclusion criteria; otherwise, we passed it to the 2^nd^ screening. For the 2^nd^ screening, we went through the full texts and filtered the articles based on the set inclusion and exclusion criteria. Only the articles accepted in the 2^nd^ screening are considered for the systematic review. As per the PRISMA guidelines, the list of the rejected articles has been provided as an appendix (see Appendix 3). Some additional articles have been selected through citation and web searching to provide a more comprehensive outlook on the topic. The screening process was conducted independently by all authors.

## Results

This section highlights the findings of the systematic review. First, we present an analytical overview for different article selection stages and later, divide the entire medical oxygen ecosystem into subtopics, addressing them in a logical sequence.

### Search result analysis

Through the screening process, we ended up with 44 articles - 41 articles found through the preliminary search and 3 additional articles appended through reference and web searching. The PRISMA selection flowchart for the study, provided in Figure 1, summarizes the selection procedure. The selection steps for the individual databases are illustrated in Table 1. The data in Table 1 contains overlaps, as some articles are present in multiple databases.

**Table 1:** Article selection statistics in different stages of screening with overlaps.

| Search Engine | Number of articles after searching | Number of articles after 1<sup>st</sup> screening | Number of articles after 2<sup>nd</sup> screening |
| --- | --- | --- | --- |
| Scopus | 136 | 82 | 26 |
| PubMed | 100 | 31 | 12 |
| SAGE | 69 | 10 | 1 |
| IEEE Xplore | 11 | 9 | 7 |
| Springer | 297 | 27 | 11 |
| Wiley | 273 | 25 | 7 |
| Other | - | - | 3 |

**Figure 1:**
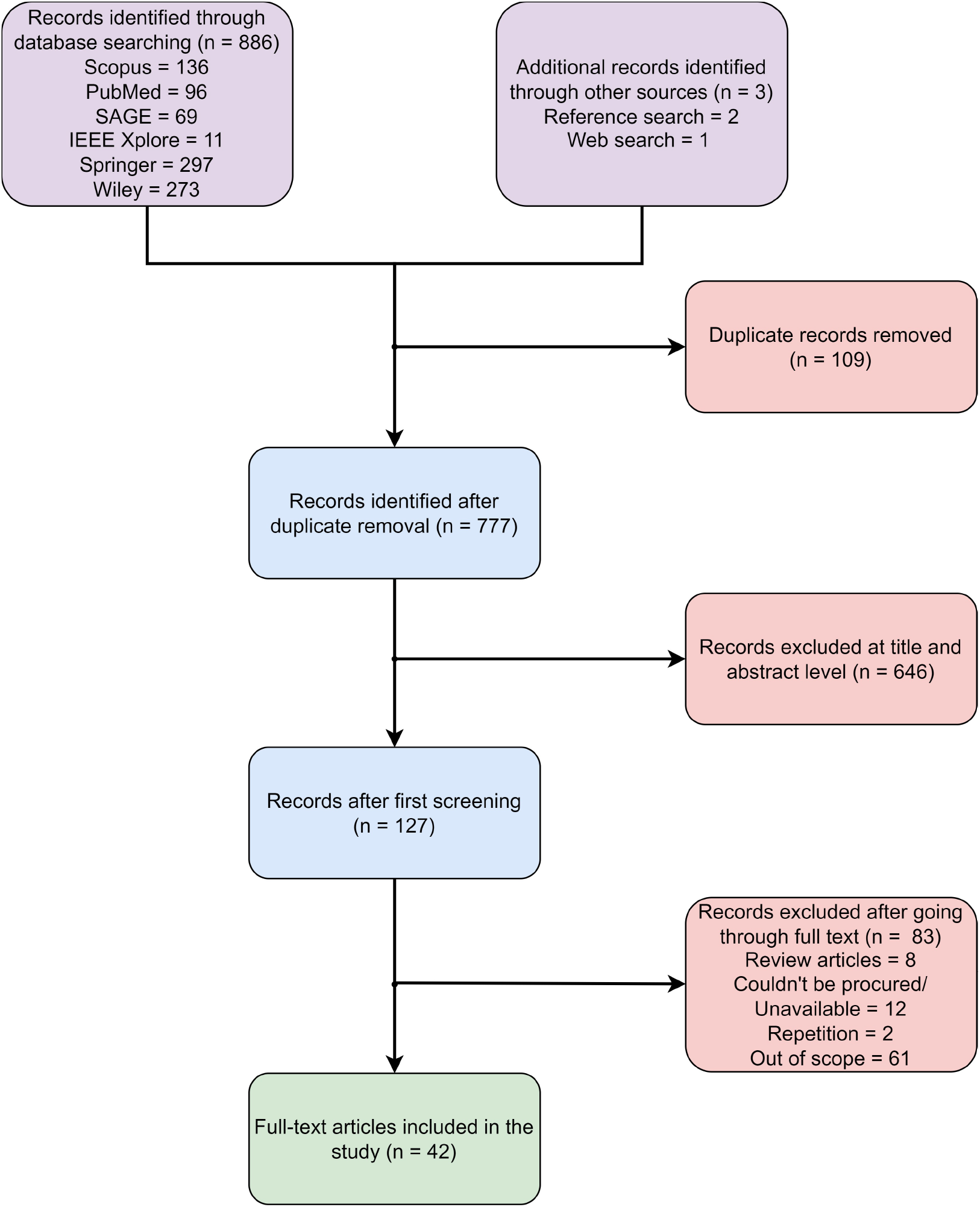
PRISMA workflow for the systematic search.

The temporal distribution of the number of selected scientific articles is presented in Figure 2, which reveals a significant lack of research prior to the COVID-19 outbreak. The sudden influx of scientific papers in 2021 aligns with WHO declaring COVID-19 as a pandemic on 11^th^ March 2020, considering the average publication delay in the domain of medicine and engineering [7, 8].

**Figure 2:**
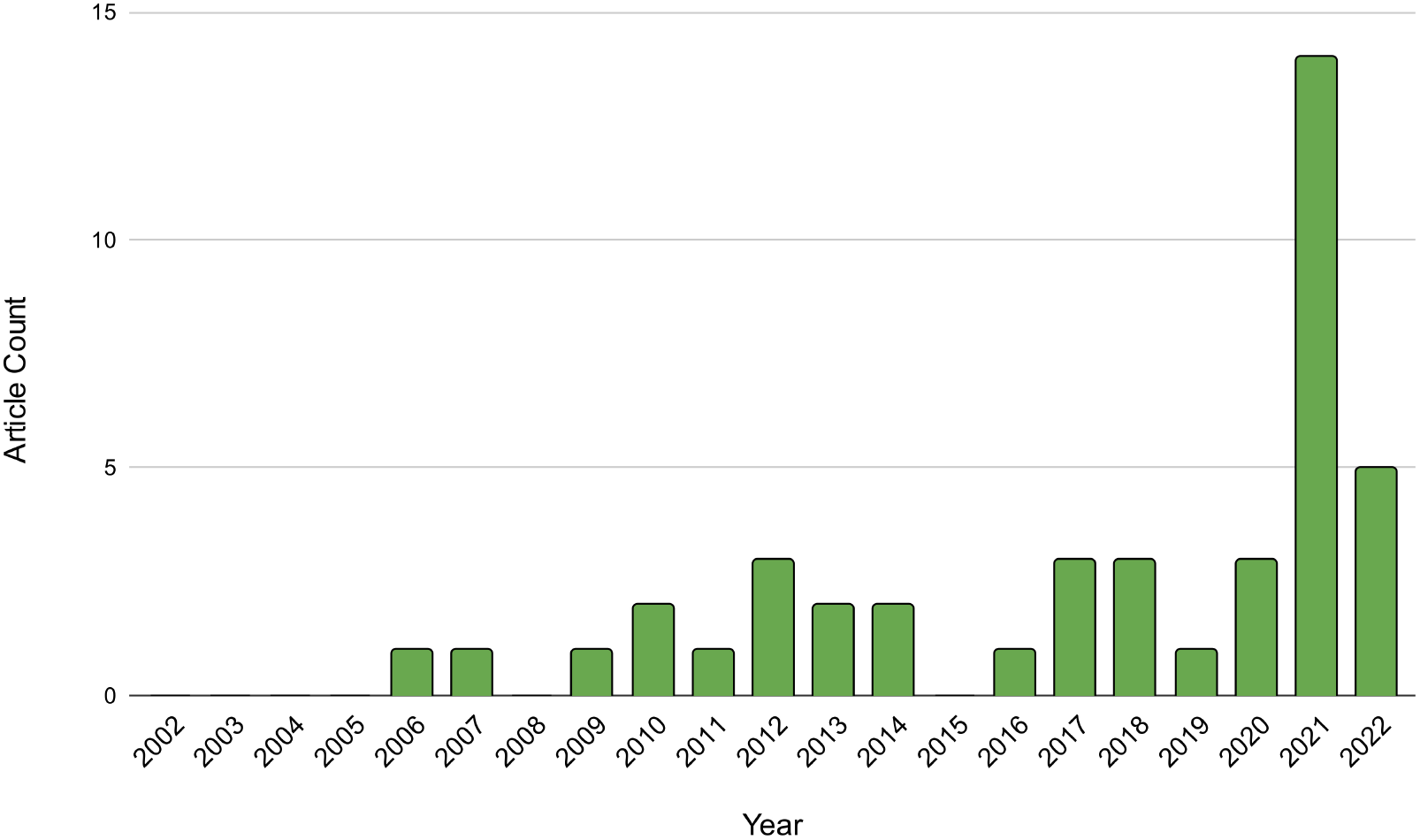
Temporal distribution of research papers in the systematic study. (The figure for 2022 is incomplete)

To further clarify the profound effect of COVID-19 on this topic, we clustered the selected articles based on the disease they targeted. Figure 3 depicts the percentage distribution of these clusters. More than half the studies (51.2%) targeted at least one specific disease. Among the targeted class, studies primarily targeting COVID-19 occupy staggering 57.2% cases, which is 28.7% more than childhood pneumonia, the highest-studied disease in this field prior to the pandemic. It goes to show how COVID-19 has affected this field.

**Figure 3:**
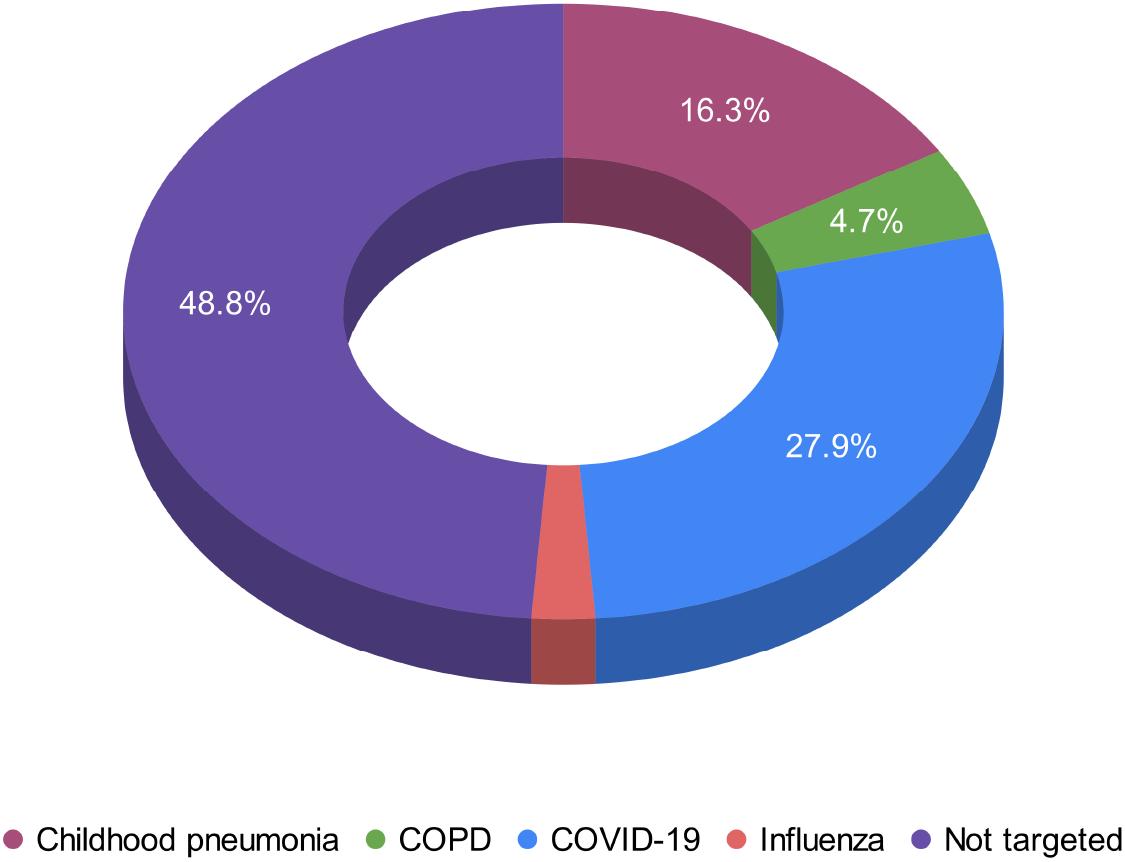
Distribution of disease targeted by selected research articles. (Only accounts for the primary interest)

### Categorizing the studies into subtopics

Since the medical oxygen ecosystem is a broad subject, we have divided it into subtopics for better discussion. For greater coherence, we have kept the categorizing in a logical order – planning, production, storage, dispensation, and monitoring. The literature distribution for each of these topics has been tabulated in Table 2.

**Table 2:** Distribution of articles across the proposed sub-categories.

| Category | Sub-category | Reference No. |
| --- | --- | --- |
| Planning | Financial analysis and planning | [9], [10], [11], [12] |
|  | Structural planning | [13], [14], [15] |
|  | Supply chain modelling and optimization | [16], [17], [18], [19], [20], [21] |
|  | Safety and protection | [22], [23], [24], [25], [26] |
| Production | Cryogenic separation | [27] |
|  | Adsorption methods | [28], [29], [30], [31], [32], [33], [34], [35], [36], [37] |
|  | Electrochemical reaction | [38], [12] |
|  | Chemical reaction | [39] |
| Storage | Cryogenic liquid | [40] |
|  | Compressed gas | [41], [42], [43], [44] |
|  | Others | [45] |
| Dispensation | Passive delivery | [46] |
|  | Active delivery | [47] |
|  | Delivery optimization | [48], [49] |
| Monitoring | Device monitoring | [50], [19] |
|  | Patient monitoring | [51], [52] |

### Planning

The first step to the establishment of a medical oxygen ecosystem is planning, both structural and financial. Although the Oxygen System Planning Tool (OSPT) by UNICEF has gained traction as a standard planning tool for oxygen systems, we believe it is vital to delve into the individual parts of the subject to address the finer elements in the field [53, 54]. As such, we have partitioned the whole planning section into four subsections - financial planning and analysis, structural planning, supply chain modeling and optimization, and safety and protection, and discussed the related articles under each subsection.

#### Financial analysis and planning

Financial analysis is generally conducted at first while setting up an oxygen system which dictates the type and scale of the system. The primary objective of financial planning is to reduce the cost of establishing a medical oxygen ecosystem, from generation to distribution, to the greatest extent feasible. As such, this section contains all articles pertaining to the cost analysis and cost-effectiveness of any component of a medical oxygen ecosystem. A total of four articles have met these criteria.

Of the four selected studies two are comparative in nature, evaluating two or more alternatives. In 2012, Bradley, Qu, Peel et al. performed a case study on The Gambia to determine whether storing electrical energy for later use by concentrators is a better option compared to storing oxygen in cylinders, when power is available [9]. Their study concluded that storing energy in batteries provides better cost optimization. Later that year, Bradley, Qu, Cheng et al. conducted an in-depth comparative analysis between five different oxygen supply technologies, considering the cost and other critical success factors [10]. The authors found the grid-charged battery-backed concentrator system to be the most effective option, followed by the low-pressure commercial generation and storage system, which is consistent with the previous study.

Unlike the comparative studies, McAllister et al. presented a study seeking to determine the optimal ratio of oxygen concentrators to medical oxygen cylinders in Fiji [11]. They centered their research on three medical centers and extended it to obtain the cost analysis for all sub-divisional hospitals nationwide. The standard in Fiji at the time of the study was to supply medical oxygen only via cylinders. The investigative study revealed that having a minimum number of concentrators with cylinder backup cost only 55% of the average status quo cost. Finally, in the fourth article, Ding et al. performed a feasibility study on Taizhou medical city to prove the cost-effectiveness of their proposed joint oxygen production in an integrated energy system (IES) model with pre-existing power-to-hydrogen (P2H) integration [12]. According to their analysis, the authors observed the status quo of purchasing O_2_ separately while keeping the IES operational to be 1,284,614 yuan (US $186,176), while producing O_2_ jointly with H_2_ costs only 278,932 yuan (US $40,425), given the extra oxygen after usage is sold off.

#### Structural planning

Medical oxygen ecosystems can vary significantly in terms of structure. While big hospitals use a central oxygen distribution system with big cryogenic reservoirs and pipes connecting the supply to the patient beds, smaller hospitals may choose to solely depend on oxygen cylinders for individual beds. The complete design is very broad and case specific and hence there is no straight-forward checklist for this type of planning to the best of our knowledge. As such, we have categorized any article that works with at least one factor associated with medical oxygen structure under this section. We found three articles meeting the required criteria.

As part of disaster management, Little et al. developed an oxygen distribution system from liquid oxygen (LOX) dewar and connected it to commercially accessible low-cost supplies [13]. The final system had been designed to supply 30 patients with up to 6 L/min of oxygen, and it cost approximately US $2,100. This study provides a holistic approach to complete structural planning using an unconventional dewar-based reservoir. The second article, dated 2021, is authored by Chen et al., who worked on developing an empirical formula to calculate the required source gas flow for a central distribution system [14]. The authors based their equation on references from [55], which provide different forms of oxygen therapy for COVID-19 patients. They verified their empirical model by comparing it to actual data collected from two novel coronary pneumonia hospitals in Wuhan, China - the Wuhan Jinyintan Hospital and the Wuhan Fire God Mountain Hospital. Although innovative, their results have discrepancies and manual calculation using their data reveals caveats in their findings. A similar study was done in the same year by Radhakrishnan et al., proposing a hypothetical model to calculate the approximate oxygen requirement for a hospital [15]. Their study considered the bed strengths and capacities of critical care units for the calculation.

#### Supply chain modelling and optimization

A significant section of the medical oxygen ecosystem is included in the supply chain, which encompasses all stages of production and delivery. Optimization of the supply chain is critical to the functioning of just about every enterprise. For this section, we have chosen all the articles involving any part of supply chain management, whether it’s delivery time or floor planning for better optimization. There are seven articles that satisfied such criteria.

The first type of article that we come across in this field tries to model the demand forecast of oxygen in some specific scenario. The earliest study in this field is from 2013 by Shen et al. performing an empirical analysis of yearly medical oxygen consumption as a case study to prove the efficacy of the grey prediction technique [16]. Their study used data collected from the First Affiliated Hospital of Shantou University Medical College from 2006 to 2012. Later in 2014, Bradley et al. performed a similar study presenting the effectiveness of the discrete event simulation (DES) model for modeling the demand for a seasonal health commodity. As a case study, they estimated the demand for oxygen for childhood pneumonia [17]. The final selected article focusing on oxygen demand forecasting comes from Saadaatamand et al. in 2022, where they predicted the necessity of oxygen-based care in the initial phases of COVID-19 using five machine learning algorithms [18]. For their study, they collected data from 398 patients from two hospitals in the Kerman province of Iran over six months.

The next article we found focuses on the delivery optimization of oxygen cylinders. This article by Carchiolo et al. tried to streamline the supply of oxygen cylinders to consumers through a homecare oxygen service platform (HOSP) [19]. The authors procured real-time data from consumers and delivery vehicles and optimized the delivery schedule using a routing algorithm. With a similar goal, Ghaithan et al. tried to develop a data-driven model to forecast oxygen gas cylinder (OGC) delivery schedules amid the soaring demand during the COVID-19 pandemic [20]. The authors performed an extensive field study by first identifying the primary elements affecting delivery time from the data taken from an OGC distribution company. The authors then developed a dataset to train their proposed machine-learning models.

The final article under this category focused on the development of crowdsourcing technology. El Barachi et al. proposed a participatory strategy to secure emergency oxygen cylinder supply for COPD patients [21]. The authors developed a geolocation-aware Android application to connect COPD or asthma patients with nearby suppliers. These suppliers could be registered service providers, private clinics, or even end-user volunteers who agreed to distribute oxygen cylinders. The authors’ conducted several validation tests of the platform’s components to guarantee targeted efficacy.

#### Safety and protection

We have opted for the incorporation of safety and protection in planning because the majority of safety analysis is completed prior to the establishment of the medical oxygen ecosystem. The other aspect of protection is the active monitoring of the whole ecosystem to detect atypical system functionality. As a result, any article focusing on risk modeling or hazard monitoring has been featured in this section.

The earliest published article under this section is by Deleris et al., which presented a quantitative model based on system analysis and probability using the probabilistic risk analysis (PRA) technique for Stanford Hospital [22]. The authors tried to fulfill the Joint Commission on Accreditation of Healthcare Organizations (JCAHO) requirements using PRA instead of the recommended failures mode and effect analysis (FMEA), arguing that FMEA could not offer transparent and comparable safety scores. A similar suggestion-based study for preliminary hazard analysis was conducted by Shaban et al., advocating the use of systems theoretical process analysis (STPA) to evaluate the risks associated with the medical gas ecosystem and oxygen delivery systems in LMICs [23]. The authors performed a case study on a hospital in Egypt to prove the model’s efficacy. Through rigorous discussion with the safety board of the hospital following the STPA framework, the authors found 13 significant hazard scenarios, as opposed to 4 scenarios found through the traditional method, illustrating the potency of this analytical strategy. Liu et al., on the other hand, introduced a data-oriented convolution weight analysis model (CWAM) to derive a new index for detecting irregular medical oxygen pipeline operation [24].

The final two articles under this section deal with fire safety. Kelly et al. report a fire breakout in the intensive care unit (ICU) by an oxygen cylinder at the Royal United Hospital (RUH), Bath, UK [25]. Based on the event, they presented various suggestions and best practices for administering medical gases. The other article by Davis et al. tried to determine the relationship between the ambient oxygen concentration and flow rate with the incidence of intra-oral fire in surgical conditions [26]. Although this study focuses on dentistry, we believe the findings are also essential for hazard analysis of regular oxygen therapy. The study found a positive, almost exponential, correlation between oxygen concentration and ignition with the minimum ignition threshold at ∼60% concentration. Although the flow rate did not affect the incidence rate, it was found to be positively correlated to the latency period of ignition.

### Production

Medical oxygen can be manufactured by a multitude of procedures, including cryogenic distillation, adsorption processes, and electrochemical and chemical reactions.

#### Cryogenic distillation

Cryogenic distillation is the standard procedure of producing a large quantity of pure oxygen in the form of liquid medical oxygen (LMO) [56]. The process is based on the Hampson-Linde cycle, which William Hampson and Carl von Linde independently patented in 1895. [57, 58]. This type of oxygen manufacturing is predominantly utilized in the metallurgical and chemical sectors, accounting for approximately 55% and 25% of utilization, respectively [59]. Tesch et al. presented a detailed comparative evaluation process of air separation units (ASUs) from both economic and exergetic point of views [27]. They described the detailed generation process in their article. Although there are variations, the general process starts by pre-treating the air and compressing it in a multi-stage compressor with inter-stage cooling. Next, a treatment process is performed when this condensed air is passed through an adsorption layer to remove the impurities like carbon dioxide, water vapour and hydrocarbons. The treated air is then passed to the fractional distillation chambers, where it is separated into three components: nitrogen, oxygen and argon. Finally, in a high-pressure fractionating chamber, the air components are separated and the liquefied oxygen is collected from the bottom of the chamber, while the nitrogen and argon rise up as vapours to the top of the chamber. The liquefied oxygen is further distilled and at the end, a purity of ∼99.5% is obtained, which can be increased further by incorporating additional distillation layers.

#### Adsorption methods

The highest studied process of medical oxygen generation according to our systematic search is the adsorption method. Some parameters are used to evaluate the performance of the generation process. The most important one is purity and it is defined as:

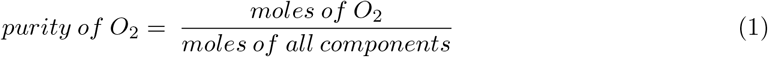

Another evaluation parameter is the recovery rate. Recovery rate is ratio of amount of O_2_ in the output to the input. The equation:

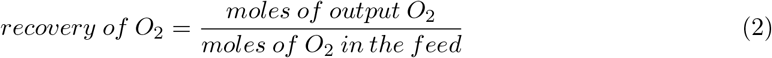

Bed size factor (BSF) relates the amount of adsorbent used to the amount of oxygen generated. It is defined by the equation:

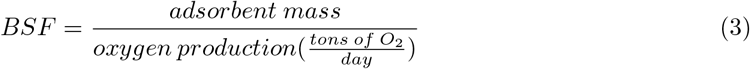

Finally, to incorporate the time function, a new parameter productivity is used and it is defined as follows:

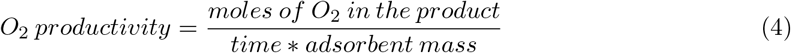

For the adsorption method of oxygen generation, different variations of the Skarstrom pulses-swing adsorption (PSA) cycle are predominantly employed [60]. It is a 4-step cycle and the steps are:

1. Compressed air is passed over a packed column zeolite at a super-ambient adsorption pressure (P_A_) producing O_2_ enriched effluent gas through the selective adsorption of N_2_
2. The zeolite column is counter-current depressurized to a near ambient final desorption pressure level (P_D_)
3. Counter-current back purge is performed on the zeolite column at P_D_ with a part of the O_2_ enriched product gas and
4. Repressurization of the column from P_D_ to P_A_ using fresh compressed air in co-current method or a part of the O_2_ enriched product gas in counter-current method or using a combination of both.

These steps are repeated to generate a continuous flow of oxygen. Throughout the literature, researchers have modified PSA in different ways to improve some specifications of their medical oxygen concentrators (MOCs). Some of the goals observed throughout the literature in terms of designing an improved MOC are – miniaturization of the device, increasing recovery rate, minimizing BSF, improving productivity etc.

In one study, authors (Rao, Farooq, and Krantz) used two-step pulsed pressure-swing adsorption (PPSA) instead of the conventional four-step PSA in order to minimize the bed size of their device, toning it down to only 0.7 cm [28]. With the same goal, a later study by Rao, Kothare, and Sircar developed a MOC using a four-step rapid pressure-swing adsorption (RPSA) cycle [29]. Tian et al. also focused on miniaturizing their six-bed MOC, but instead of using a modified version of PSA, they introduced a rotary distribution valve to save space [30]. Some of the studies have focused on developing portability on top of miniaturization. Pan et al. developed their portable MOC using the pressure vacuum swing adsorption (PVSA) cycle, while Vemula et al. used the RPSA cycle [31, 32]. The MOC developed by Vemula et al. could work as a regular unit or a standalone unit, although working as a standalone unit sacrificed performance for portability.

Some studies have focused on improving the general performance and productivity of MOCs. Zhu and Wang proposed a PSA variation - rapid vacuum pressure swing adsorption (RVPSA) with intermediate gas pressurization to improve the performance of miniature MOCs [33]. In another study by Zhu, Liu et al., the authors utilized the short cycle time of rapid cycle pressure swing adsorption (RCPSA) to increase the productivity of miniature MOCs [34]. Qadir et al. performed a different type of study, where they developed a numerical model of a two-bed MOC system using the RPSA cycle and studied its performance on three different adsorbents-Li-LSX, Na-LSX, and K-LSX [35]. They considered these adsorbents’ pellet sizes (range: 0.025-0.1 cm) for their experiment and concluded that 0.1 mm Li-LSX zeolite provided the best performance. Arora et al., in a similar study, explored the performance flexibility of the PSA and PVSA cycles with three different adsorbents - LiX, 5A and LiLSX through a numerical simulation [37]. They concluded that PVSA using LiLSX as the adsorbent gave the maximum performance flexibility with the best parameter results.

We have tabulated the specifications of all these MOCs in Table 3 for a better comparison. We have listed only the best-performing specifications in case a study included a comparative analysis. It was not possible to convert all the values to one standard unit for the lack of required information in the studies. Also, some values are missing in the table as they were not listed in the articles.

**Table 3:** Properties of the medical oxygen concentrators found in the literature. *PPSA* - pulsed pressure-swing adsorption, *RPSA* - rapid pressure-swing adsorption, *RVPSA* - rapid vacuum pressure swing adsorption, *RCPSA* - rapid cycle pressure swing adsorption, *SL* - standard liter, *TPD* - tons per day.

| Reference | Cycle used | Adsorbent used | Flow rate | Purity | Recovery | BSF | Productivity |
| --- | --- | --- | --- | --- | --- | --- | --- |
| Rao et al. [28] | PPSA | 5A, AgLiX | 5 L/min | >90% | 10-25% (5A),<br>10-55% (AgLiX) | - | - |
| Rao et al. [29] | RPSA | LiLSX | 1-3 SL/min | ~90% | 29.3% | 45 kg/TPD | - |
| Tian et al. [30] | PSA | 5A | 0.1 m<sup>3</sup>/min | >93% | >57% | - | 0.0357<br>m<sup>3</sup>·h<sup>-1</sup>·kg<sup>-1</sup> |
| Pan et al. [31] | PVSA | JLOX-101<br>LiX | 1.128 L/min | 91.64% | - | - | - |
| Vemula et al. [32] | RPSA | LiLSX | 10 SL/min<br>(regular), 5.8<br>SL/min<br>(standalone) | 90% | ~34% | ~45 kg/TPD<br>(regular),<br>~95 kg/TPD<br>(standalone) | - |
| Zhu et al. [33] | RCPSA | LiLSX | 1 L/min | ~92% | ~30% | ~78 kg/TPD | - |
| Zhu et al. [34] | RVPSA | LiLSX | 0.75 L/min | 90% | 29.45% | 82.84<br>kg/TPD | - |
| Qadir et al. [35] | RPSA | LiLSX | 1 L/min | 94% | 40% | 78 kg/TPD | 4.7<br>mmol·kg<sup>-1</sup>·s<sup>-1</sup> |
| Arora et al. [37] | PVSA | LiLSX | 1-15 L/min | 93-95.7% | - | - | - |

The final article in this field focuses on the susceptibility of MOCs to external disturbances that can hamper the continuous flow of oxygen or even lower the purity of the output. Urich et al. proposed a multivariable model predictive controller (MPC) to counter these external disturbances in an RPSA-based single-bed cyclic MOC [36]. They designed the controller to maintain a purity of 90% by dynamically controlling the duration of different RPSA steps. They performed a case study with both PID controller and multivariate MPC and observed that MPC performed better.

#### Electrochemical generation

After the adsorption-based generation, water electrolysis appeared as the most studied oxygen generation method. The problem with this method is that it produces hydrogen gas as a by-product which is a fire hazard and needs to be disposed of properly. Giddey et al. investigated the electrolytic generation of oxygen utilizing a Nafion polymer electrolyte membrane electrolyzer to eliminate the produced hydrogen automatically. Based on their investigation, they developed a device that could increase the power efficiency of electrochemical generation of oxygen by more than 35% [38]. The main goal of their study was to develop an oxygen generation system that is portable, compact, quiet, and reliable with the PSA systems set as reference. Each cell of the proposed device had an active area of only 100 cm^2^. A two-cell stack could operate up to 0.6 A/cm^2^ without any hydrogen generation while outputting 418 mL/min of pure oxygen.

The electrolysis of water is not only used to generate oxygen, but also hydrogen in specific use cases. In such scenarios. oxygen is produced as a by-product and removed. Recently IES has been gaining popularity with the integration of intermittent renewable sources, with some of the newer IES using the P2H system as an improved economical approach [61]. Ding et al. proposed an update on such an IES with an incorporated alkaline P2H system to utilize the by-product oxygen to generate medical oxygen via purification [12]. The authors reported that their novel P2H-and-Oxygen (P2HO) system can significantly aid in the oxygen crisis of COVID-19 and that, when optimized, this form of medical oxygen generation proves remarkably less expensive than the conventional cryogenic distillation technique. They have performed a test study on China Medical City, Taizhou, Jiangsu province, China to validate their findings.

#### Chemical generation

In addition to the more studied adsorption and electrochemical oxygen manufacturing processes, Dingley et al. developed a purely chemical oxygen manufacturing strategy [39]. Their main goal was to ensure a steady flow of medical oxygen during the time of emergency without the input of compressed gas. After extensive experimentation with reagent combination, the authors concluded the optimum combination to be 1 L water, 0.75 g manganese dioxide catalyst, 60 g sodium percarbonate granules and 800 g of custom pressed percarbonate pellets. Their device was able to output 99% pure oxygen in a steady flow for more than 90 minutes.

### Storage

As discussed in the production section, medical oxygen is mainly generated in liquid and gaseous forms. In this section, we discuss the storage methods for both these forms.

LMO storages are mainly of three types – dewar, cryogenic cylinders, and cryogenic storage tanks [40]. Dewars and cylinders are analogous in that they allow portability at the cost of volume. The main difference is that dewars are non-pressurized containers, whereas cylinders store high-pressurized LOX. The typical volume of commercially available dewar flask spans from 5 to 200 L, whereas conventional cryogenic cylinders can store 80 to 450 L of LOX at a pressure of around 24 atm. Cryogenic storage tanks provide large storage volumes at high pressure, and the usual volume of these reservoirs ranges from 500 to 420,000 gallons. The most straightforward configuration of a cryogenic storage tank consists of a tank, a vaporizer, and a pressure controller. Typically, they are only installed in large medical institutions. The cryogenic reservoir is attached to a distribution pipeline that spans the hospital.

On the other hand, the gaseous form of medical oxygen is mainly stored in cylinders after compression [41]. However, the literature from the systematic search has made some suggestions regarding this type of storage [42–45]. These studies have been conducted in vulnerable economic regions with intermittent power availability, and they argued the economic feasibility of oxygen cylinders. To improve the cost-effectiveness of storing oxygen, Rasool et al. developed a low-pressure storage device to replace high-pressure cylinders [42]. The device had a storage capacity of 200 L and used water as a ballast. Peake et al. conducted a three-month field trial in the Mbarara region of Uganda to prove the efficacy of this device [43]. During the entire trial, the device could maintain a steady output of 8 L/min to four pediatric beds, and it was able to save around US $1,061 compared to its status quo of backup cylinders. Similarly, Otiangala et al. deployed a medium-pressure reservoir (MPR) that could supply oxygen at 1-5 L/min as part of a clinical trial. The MPR was able to ensure continuous oxygen flow for regular power outages [44]. Bradley, Cheng, et al. took a completely different approach to the problem. They proposed the usage of MOCs with backup power storage instead of wholly relying on oxygen cylinders [45].

### Dispensation

This section covers the patient-side delivery mechanism. Oxygen administration can be divided into two categories: passive delivery, in which oxygen is provided to the patient without mechanical intervention, and active delivery, in which oxygen is mechanically vented to the patient’s lungs. We have also included delivery optimization under this section to provide a complete picture.

#### Passive delivery

There are primarily two passive oxygen delivery strategies for patients:

1. Semi-invasive nasal cannula
2. Non-invasive oxygen mask

Nasal cannulae or nasal prongs can provide oxygen efficiently at lower flow rates compared to non-invasive oxygen masks [62]. As such, oxygen masks are expensive alternatives to nasal prongs. Oxygen masks, although expensive compared to nasal cannulae, provide improvement in quality of life as they are a more comfortable alternative for the patient. Different types of oxygen masks are available, including oral-nasal masks, nasal masks, and even full-face masks, depending on the region covered. A recent development in passive oxygen delivery using masks was established by Karam et al., who proposed an aerosol barrier mask (ABM) to prevent the spread of SARS-CoV-2 from afflicted patients [46]. This mask is integrated with the option to provide therapeutic oxygen. The ABM nullifies almost all aerosolized particles and droplets originating from the patient at proximity. Additionally, the authors performed a carbon dioxide accumulation analysis on the mask and found that the mask does not allow abnormal accumulation, deviating too much from the norm.

#### Active delivery

Patients with respiratory impairment due to any form of physical implication need mechanical ventilation. Ventilators simulate the work of the lungs for the patients and push oxygen enriched air to the lungs. The air is generally dispensed in two ways [63]:

1. Non-invasive face mask for less severe cases
2. Invasive breathing tube for acute conditions

Ventilator design has become a trending topic since COVID-19 precipitated the worldwide shortage of ventilators [64]. Unfortunately, our systematic search has yielded only one ventilator design article as the topic falls under a gray area for our study. Ventilator design is not primarily associated with the medical oxygen ecosystem as it is more of an accessory to the system. The only article we found on the topic comes from the design and development team of Vikram Sarabhai Space Centre, Thiruvananthapuram, India. They have presented the complete design with prototype development and testing of three low-cost ventilators - SVASTA, PRANA, and VaU to tackle the surge of ventilator demand during the COVID-19 pandemic [47].

#### Dispensation optimization

Dispensation optimization of medical oxygen supply, in its simplest form, involves reducing unwanted consumption. With this goal in mind, Cavaglià et al. proposed a closed-circuit noninvasive (NIV) continuous positive airway pressure (CPAP) to minimize oxygen consumption, along with a decrease in air contamination by COVID-19 pathogens and noise [48]. Their proposed NIV-CPAP system included automatic control of patients’ respiratory parameters. The prototype system could limit oxygen feeding up to a substantial 30-fold compared to standard open circuit systems while maintaining the same FiO_2_ levels. Zheng et al., on the other hand, proposed an automated system based on reinforcement learning (RL) to control the flow of oxygen to patients [49]. The model reportedly decreased the mean mortality rate of COVID-19 patients by 2.57% while maintaining a lower average flow (1.28 L/min) compared to standard care procedures.

### Monitoring

Monitoring is critical to maintaining a proper working state of the overall medical oxygen system. This section discusses the findings of articles focused on monitoring any aspect of the medical oxygen ecosystem.

Maintaining a constant oxygen distribution in a medical facility is of the utmost importance. As such, it is necessary to ensure the correct working of all parts of the pipeline. Different sensors are integrated at different parts of the ecosystem to ensure proper functionality, from generation to distribution [65]. One recent work in this field comes from Muhammad et al. They proposed a low-pressure alarm device to be incorporated with medical oxygen cylinders to minimize the risk of insufficient oxygen flow to the patients [50]. Carchiolo et al., on the other hand, performed a study on monitoring domiciliary oxygen therapy proposing an improved HOSP harnessing the power of industrial IoT (IIoT) [19]. They optimized their service by collecting real-time data and updating the scheduling. The primary concerns of their study were monitoring oxygen consumption, maintenance requirements for medical equipment, oxygen amount abuse according to the treatment plan, and oxygen cylinder replacement timing.

Another crucial aspect of oxygen monitoring is patient-based blood oxygen saturation (SpO2) monitoring. It is mainly done by measuring the oxygen concentration in arterial blood. The most common method of such measurement is done through the use of pulse oximetry [66]. A recent work by Dipayan used the optical sensor MAX30102, which has an integrated pulse oximeter to measure both SpO2 and the heart rate of a patient [51]. The aim of the study was to develop an internet of things (IoT) based patient vital signs tracking. Gunasekar et al. also proposed an IoT-based patient monitoring device. Instead of measuring the SpO2 of the patient, it measured the oxygen level in the cylinder. For the measurement, authors used a load cell with 80 kg calibrated using the HX 711 module [52].

## Discussion

In this section we discuss on the research gaps we found throughout our systematic review, and present some recommendations where applicable:

1. **Standardizing checklists for structural planning:** Through our systematic search, we found three research articles on structural planning, but none focused on standardizing or improving the checklists for structural planning. We acknowledge that it may not be possible to work towards a generalized solution as medical oxygen systems vary drastically from one healthcare facility to another. We recommend classifying oxygen systems into standard categories in such cases and developing individual checklists for each.
2. **Supply chain optimization:** Although we have six scientific articles on supply chain optimization, a significant portion of the total number of accepted articles, the studies have overlooked several crucial aspects. We have found no work that tries to model the oxygen demand in a medical facility. Shen et al. have come close to the field by performing an empirical study to determine the annual oxygen consumption using the grey prediction model [16]. However, we need short-term, real-time models to help healthcare facilities plan and prepare ahead of time. Short-term demand forecasting will also allow the conservation of oxygen by maximizing optimization.
3. **Crisis management and planning:** Throughout the systematic review, we found only one article that came close to this field of study. The study was conducted by Little et al. and focused on developing an improvised oxygen pipeline for a medical facility in case of a disaster scenario [13]. It was quite perplexing to see no other article remotely focusing on this topic. We speculate that this lack of relevant studies in this field contributed to the widespread haphazardness during the COVID-19 pandemic. We would encourage researchers to work in this area so that we can be better prepared in case there is a similar disaster in the future.
4. **Study the chemical generation method of medical oxygen:** Although not conventional, we have found one promising paper by Dingley et al. on a purely chemical method of generation of medical grade oxygen for emergencies [36]. Further research can be performed in this field to make the generation more sustainable as they can prove very useful in any disaster scenario.
5. **Integration of optimized energy storage to improve concentrator deployment:** Bradley et al. verified the cost-effectiveness of energy storage compared to compressed gas storage [9, 10]. The last work in this field was published in 2012, with the field seeing no more development after that. Large-scale integration of such a setup with further optimization can be a great field of research.
6. **Automated dispensation optimization:** Dispensation optimization is a vital field that had no work until very recently. Zheng et al. tried to improve the effectiveness of oxygen therapy while mitigating unwanted loss using the RL algorithm [49]. This type of automated control for oxygen therapy is new and should get more attention, at least in this age of technology.

## Conclusion

Medical-grade oxygen is one of the most widely utilized medicines in the world today. During the COVID-19 outbreak, the necessity of oxygen management has been acknowledged globally, which has engendered a recent upsurge of research on this topic. Our work aimed to offer a comprehensive outlook on the current state of the subject matter. As such, we have performed a systematic assessment following the PRISMA protocol, spanning the past 20 years.

Due to the paucity of current research on this topic, it has been challenging to undertake a systematic review. Still, we have sought to present a complete image of this field by combining a multitude of complementary articles. As part of our analysis, we have outlined several research gaps and potential future developments in this topic. We expect that this article will serve as an asset for researchers in the domain of the clinical oxygen ecosystem, endorsing future efforts.

## Limitations of study

We wish to clarify that our attempt to encompass an extensive scope of study may have resulted in certain shortcomings in specific fields, such as the ventilator design section. Because this is a systematic review, we have avoided including information from our part without adequate referencing. Even so, certain sections have necessitated the addition of extra articles to offer some insight into inevitable topics such as cryogenic distillation of oxygen generation, which, although being the primary method of production, have not yielded any research papers during our systematic search. In such instances, we have attempted to keep the content as concise as possible to avoid any information bias.

## Supporting information

Appendix 1 PRISMA Checklist

Appendix 2 Search Strings

Appendix 3 Exclusion reasoning

## Data Availability

All data produced in the present work are contained in the manuscript

## Acknowledgements

The authors would like to thank Dr. M Mostafa Zaman for providing his knowledgeable insights and informed advice.

### Appendix 1: PRISMA-P checklist

The PRISMA-P checklist has been provided as additional file named “Appendix 1: PRISMA Checklist”

### Appendix 2: Complete search strategy across different engines

The exact search strings used in different search engines have been listed under the file “Appendix 2: Search Strings”

### Appendix 3: List of articles excluded during second screening with reasoning

The exclusion reasoning has been provided under the file “Appendix 3: Exclusion Reasoning”

