## Appendix 2 Search Strings for "A Systematic Review on Medical Oxygen Ecosystem: Current State and Recent Advancements"

The last search was made on 15th May 2022. The search engines are susceptible to updates afterwards and can affect the number of articles found through search. We reported the number of articles we found during our searches. Our detailed search strings and techniques have been provided in this document.

**Scopus string:**

TITLE-ABS-KEY ( ( "medical oxygen" OR "hospital oxygen" OR "clinical oxygen" ) AND ( manag* OR optim* OR forecast* OR ( demand AND forecast* ) OR predict* OR ( demand AND predict* ) ) ) AND ( ( PUBYEAR = 2022 OR PUBYEAR = 2021 OR PUBYEAR = 2020 OR PUBYEAR = 2019 OR PUBYEAR = 2018 OR PUBYEAR = 2017 OR PUBYEAR = 2016 OR PUBYEAR = 2015 OR PUBYEAR = 2014 OR PUBYEAR = 2013 OR PUBYEAR = 2012 OR PUBYEAR = 2011 OR PUBYEAR = 2010 OR PUBYEAR = 2009 OR PUBYEAR = 2008 OR PUBYEAR = 2007 OR PUBYEAR = 2006 OR PUBYEAR = 2005 OR PUBYEAR = 2004 OR PUBYEAR = 2003 OR PUBYEAR = 2002 ) AND ( LANGUAGE ( english ) ) ) AND ( LIMIT-TO ( SRCTYPE , "j" ) OR LIMIT-TO ( SRCTYPE , "p" ) )

- Manual search was done to remove articles before November 2002

**PubMed string:**

("medical oxygen" OR "hospital oxygen" OR "clinical oxygen") AND ( manag* OR optim* OR forecast* OR ( demand AND forecast* ) OR predict* OR ( demand AND predict* ) )

- 2002 November

**Sage:**

("medical oxygen" OR "hospital oxygen" OR "clinical oxygen") AND ( manag* OR optim* OR forecast* OR ( demand AND forecast* ) OR predict* OR ( demand AND predict* ) )

- From 2002
- Manual search was done to remove papers before November of 2002

**IEEE:**

((medical oxygen) OR (hospital oxygen) OR (clinical oxygen)) AND (manag* OR optim* OR forecast* OR (demand forecast*) OR predict* OR (demand predict*))

- Conferences and Journals
- Since 2002

**Springer:**

("medical oxygen" OR "hospital oxygen" OR "clinical oxygen") AND (manag* OR optim* OR forecast* OR (demand forecast*) OR predict* OR (demand predict*))

- Articles
- Conference Paper
- English
- 2002 onwards
- Manual search was done to remove papers before November of 2002

**Wiley:**

(("medical oxygen") OR ("hospital oxygen") OR ("clinical oxygen")) AND ((manag*) OR (optim*) OR (forecast*) OR (demand forecast*) OR (predict*) OR (demand predict*))" anywhere

- Journals (Excluded books and referenced works)
- November 2002 onwards
