## Appendix 3 Exclusion reasoning for "A Systematic Review on Medical Oxygen Ecosystem: Current State and Recent Advancements"

### Articles excluded in the 2nd screening

#### Repetition

These articles have revised elaborate versions available. There are only two articles meeting this criteria - [1], [2]

#### Unable to be procured/ Unavailable

Some of these articles couldn't be procured for lack of access of the authors and the others were unavailable. The twelve articles meeting these criteria are - [3], [4], [5], [6], [7], [8], [9], [10], [11], [12], [13], [14]

#### Review articles

Although the searches were made to exclude review papers, some review papers made it through. There are a total of eight review papers found through the database searching. These are
- [15], [16], [17], [18], [19], [20], [21], [22]

#### Out of scope

These section consists of articles that didn't meet all the inclusion criteria. Some were excluded because their article type didn't meet the inclusion article type, some were excluded because they focused on a tertiary topic with respect to the main issue. There are a few articles that studied some hypothetical scenarios and didn't go into the real world implications. All these have been grouped under this section. There are a total of sixty-one articles. They are
- [23], [24], [25], [26], [27], [28], [29], [30], [31], [32], [33], [34], [35], [36], [37], [38], [39], [40], [41], [42], [43], [44], [45], [46], [47], [48], [49], [50], [51], [52], [53], [54], [55], [56], [57], [58], [59], [60], [61], [62], [63], [64], [65], [66], [67], [68], [69], [70], [71], [72], [73], [74], [75], [76], [77], [78], [79], [80], [81], [82], [83]

17. Jha M, Gaur N. Life cycle of medical oxygen from production to consumption. Journal of family medicine and primary care;11:1231–1236.

18. Finer N, Leone T. Oxygen saturation monitoring for the preterm infant: the evidence basis for current practice. Pediatric research;65:375–80.

19. O'Driscoll BR, Smith R. Oxygen Use in Critical Illness. Respiratory care;64:1293–1307.

20. Davidescu AA, Apostu SA, Stanciu-Mandruleanu C. Shedding Light on the Main Characteristics and Perspectives of Romanian Medicinal Oxygen Market. Healthcare. 2021;9(2). doi:10.3390/healthcare9020155.

21. Severinghaus JW. The history of clinical oxygen monitoring. International Congress Series. 2002;1242:115–120. doi:10.1016/S0531-5131(02)00723-9.

22. Ismail J, Bansal A. Medical Oxygen: A Lifesaving Drug During the COVID-19 Pandemic–Source and Distribution. Indian Journal of Pediatrics;89(6):607–615. doi:10.1007/s12098-021-03978-0.

29. Shen G, Wu X, Tang C, Yan Y, Liu J, Guo W, et al. An oxygen enrichment device for lowlanders ascending to high altitude. BioMedical Engineering OnLine;12(1):100. doi:10.1186/1475-925X-12-100.

39. Fu C, Caruso P, Lucatto JJJ, de Paula Schettino GP, de Souza R, Carvalho CRR. Comparison of two flow generators with a noninvasive ventilator to deliver continuous positive airway pressure: a test lung study. Intensive Care Medicine;31(11):1587–1592. doi:10.1007/s00134-005-2795-x.

40. Tolla HS, Woyessa DB, Balkew RB, Asemere YA, Fekadu ZF, Belete AB, et al. Decentralizing oxygen availability and use at primary care level for children under-five with severe pneumonia, at 12 Health Centers in Ethiopia: a pre-post non-experimental study. BMC Health Services Research;22(1):676. doi:10.1186/s12913-022-08003-4.

41. Considine J, Botti M, Thomas S. Descriptive analysis of emergency department oxygen use in acute exacerbation of chronic obstructive pulmonary disease. Internal Medicine Journal. 2012;42(4):e38–e47. doi:10.1111/j.1445-5994.2010.02220.x.

42. Gurkin VN, Kagramanov GG, Loiko AV, Farnosova EN, Blanko-Pedrekhon AM, Milyaev AV. Development of a Portable Membrane Oxygen Concentrator. Membranes and Membrane Technologies. 2021;doi:10.1134/s2517751621030045.

43. Huang Y, Mian Q, Conradi N, Opoka RO, Conroy AL, Namasopo S, et al. Estimated Cost-effectiveness of Solar-Powered Oxygen Delivery for Pneumonia in Young Children in Low-Resource Settings. JAMA Network Open. 2021;4(6):e2114686–e2114686. doi:10.1001/jamanetworkopen.2021.14686.

44. Rama Rao V, Chai SW, Kothare MV, Sircar S. Highlights of non-equilibrium, non-isobaric, non-isothermal desorption of nitrogen from a LiX zeolite column by rapid pressure reduction and rapid purge by oxygen. Adsorption;20(2):477–481. doi:10.1007/s10450-013-9581-9.

45. Lake D, Henwood M, Eadie P, Dagg K, Searle P, Fenlon S, et al. How not to run out of oxygen: Primary FRCA forcibly put into practice by COVID-19 to maintain patient safety. Journal of Patient Safety and Risk Management;26(2):81–88. doi:10.1177/2516043521996580.

52. Kumar A, Gunnam PR, Jat B, Kant R. Medical gas pipeline system as a limiting factor for hospital oxygen supply during COVID-19 outbreak. Indian Journal of Anaesthesia;65(11).

53. Bikkina S, Kittu Manda V, Adinarayana Rao UV. Medical Oxygen Supply During COVID-19: A Study with Specific Reference to State of Andhra Pradesh, India. Materials today Proceedings;doi:10.1016/j.matpr.2021.01.196.

54. DeCoste JB, Weston MH, Fuller PE, Tovar TM, Peterson GW, LeVan MD, et al. Metal–Organic Frameworks for Oxygen Storage. Angewandte Chemie International Edition. 2014;53(51):14092–14095. doi:10.1002/anie.201408464.

55. Weismann D, Reißmann H, Maisch S, Füllekrug B, Schulte J. Monitoring of Functional Residual Capacity by an Oxygen Washin/Washout; Technical Description and Evaluation. Journal of Clinical Monitoring and Computing;20(4):251–260. doi:10.1007/s10877-006-9029-9.

56. Rao VR, Kothare MV, Sircar S. Numerical simulation of rapid pressurization and depressurization of a zeolite column using nitrogen. Adsorption;20(1):53–60. doi:10.1007/s10450-013-9548-x.

57. Hangan H, Bekele S. Numerical simulation of the flow and concentration fields for oxygen delivery systems. Journal of biomechanical engineering;125:648–62. doi:10.1115/1.1613296.

65. Pollock NW, Natoli MJ. Performance characteristics of the second-generation remote emergency medical oxygen closed-circuit rebreather. Wilderness environmental medicine;18:86–94.

72. Conradi N, Mian Q, Namasopo S, Conroy AL, Hermann LL, Olaro C, et al. Solar-powered oxygen delivery for the treatment of children with hypoxemia: protocol for a cluster-randomized stepped-wedge controlled trial in Uganda. Trials;20(1):679. doi:10.1186/s13063-019-3752-2.

79. Duprez F, Dubois A, Ollieuz S, Cuvelier G, Reychler G. Thorpe tube and oxygen flow restrictor: what's flow accuracy? Journal of Clinical Monitoring and Computing;35(2):337–341. doi:10.1007/s10877-020-00476-0.

80. Satpathy SK, Upadhyay MR, Upadhyay R, Dash MK, Nayak AS, Naik S. Use of Low-Flow Flowmeter to Titrate the FiO2 Delivered in Neonatal Resuscitation. Journal of Neonatology;35(3):104–107. doi:10.1177/09732179211040619.

81. Wong MD, Chung H, Chawla J. Using continuous overnight pulse oximetry to guide home oxygen therapy in chronic neonatal lung disease. Journal of Paediatrics and Child Health. 2020;56(2):309–316. doi:10.1111/jpc.14606.

82. Madzimbamuto FD. Ventilators are not the answer in Africa. African Journal of Primary Health Care and Family Medicine. 2020;12(1). doi:10.4102/PHCFM.V12I1.2517.

83. Thorleuchter D, Van Den Poel D. Weak signal identification with semantic web mining. Expert Systems with Applications. 2013;40(12):4978 – 4985. doi:10.1016/j.eswa.2013.03.002.
